# Nearly Perfect Forecasting of the Total COVID-19 Cases in India: A Numerical Approach

**DOI:** 10.1101/2020.06.13.20130096

**Authors:** Hemanta K. Baruah

## Abstract

There are standard computational and statistical techniques of forecasting the spread pattern of a pandemic. In this article, we are going to show how close the forecasts can be if we use a simple numerical approach that can be worked out using just a scientific calculator. Using a few recent data, short term forecasts can be found very easily. In this numerical technique, we need not make any assumptions, unlike in the cases of using computational and statistical methods. Such numerical forecasts would be nearly perfect unless the pandemic suddenly starts retarding during the period of the forecasts naturally or otherwise.

## Introduction

Quite a few computer dependent mathematical models to forecast the pattern of spread of infectious diseases are available in the literature. Models of this type are constructed under certain assumptions and are defined in terms of simultaneous differential equations. Meyers [1] studied about forecasting of the spread in the case of small epidemics, using the Susceptible-Infected-Recovered (SIR) model. Wu *et. al*. [2] used the Susceptible-Exposed-Infectious-Recovered (SEIR) model in a simulation study of the COVID-19 spread. Anastassopoulou *et. al*. [3] used the Susceptible-Infectious-Recovered-Dead (SIRD) model in a simulation study of the COVID-19 situation in the very initial stage of the pandemic. Ghosh *et. al*. [4] used the Susceptible-Infectious-Susceptible (SIS) pandemic model to forecast COVID-19 spread in India. It may be noted that to execute such models one must get precise data regarding the parameters concerned. Further, which particular model is the best is not yet clear.

Another approach to forecast the spread of a pandemic is time series analysis. Standard procedures such as auto-regressive integrated moving average (ARIMA) are there in time series analysis, and using these procedures forecasts can be made. When a time series is analyzed, forecasts are made with reference to upper and lower limits expressed in terms of certain probability level of confidence. Kumar *et. al*. [5] have used the ARIMA method of time series analysis to study the Indian situation. Poonia and Azad [6] and Azad and Poonia [7] have studied the problem in two phases using the ARIMA method. Their forecasts were quite close to the actual values observed later. Basu [8] studied time dependent spread of the virus in India using his own model. As per his forecasts expressed in the form of a graph, the total number of cases in India should cross 200,000 in the beginning of June, and his forecast has been found to be true.

In this article, we are going to discuss how short term forecasts can be made based on a simple numerical analytical approach without using probabilistic standpoints and without using pandemic models. We shall show that this approach works nearly accurately in the case of short term forecasts. We have observed that this kind of a situation can arise in the case of studying the spread pattern of a pandemic of the type of COVID-19. Our approach may give us a slightly overestimated forecast, but they can be useful for the policy makers for immediate decisions. For our discussions, we shall consider the current spread pattern in India.

The COVID-19 pandemic has spread the world over since the beginning of the current year. It has been observed that in certain countries such as Italy, United Kingdom and Spain, the spread pattern was an increasing function, increasing in a very highly nonlinear manner, and after some points of time the growth had started to subside. In countries such as the United States and Russia, it is still increasing in a highly nonlinear manner. In India also it is still increasing highly nonlinearly. Therefore if an attempt is made to forecast the spread pattern taking the world as a whole, the forecasts would be overestimations, because within the period of the forecast the growth pattern may slow down in some country making the forecasts overestimations thereby. If we consider forecasting of the spread pattern not in the world as a whole but in an individual country, then also the forecast may actually be an overestimate. In the United States, the spread was very nearly exponential till May 8, but from May 9 onwards for a few days the growth subsided, and therefore forecasts of the spread pattern for those few days would have resulted in overestimations.

In India, it was nonlinear right from the beginning. Currently the pattern is nearly exponential. How long it would continue to be so is uncertain. The spread may hopefully subside suddenly or it may continue to grow still further. Therefore forecasting of the spread pattern in India can be made for short terms only.

In what follows, we are going to discuss a procedure of looking at the time series data upside down, and establish a simple numerical model based on some recent values, which in turn can be successfully utilized for short term forecasting. After comparing with the observed values, if we see that the forecasts are quite acceptable, then that should certify that our numerical approach does work well.

## Methodology

Standard methods available in the statistical literature can deal with time series data for rigorous stochastic analysis that can ultimately lead to forecasts with reference to some prefixed probability level of confidence. Methods such as auto regressive integrated moving average have been used to find forecasts of the COVID-19 spread in India. Models used for infectious diseases have also been tried upon. In a few cases the forecasts made in the very initial stage of the outbreak did result in very good forecasts, but in some cases the forecasts were not quite appropriate. It is true that statistical randomness is inherent in the data concerned. Therefore statistical analysis is the most appropriate for studying this kind of data. At the initial stage of the pandemic, randomness did play a major part. However, as time passed the total number of cases started to grow in such a fast pace that in comparison to the mathematical part, the random part in the time series slowly started to become less and less important. Therefore a numerical approach to deal with forecasting may be sufficient. There may not be any mathematical rigor involved in the technique, but still short term forecasts can be made after observing the recent data taking the time series upside down. This is our working hypothesis, and we have seen that our hypothesis has worked well in the Indian case.

Let *N*(*t*) be the total number of cases at a given time *t*. The total number of cases includes the active cases, the recovered cases and the number of deaths. If we observe the graphical representation carefully, it is not difficult to surmise what kind of mathematical form the data are following. We have used the COVID-19 data published by Worldometers.info [9]. The data are available in the form of graphs and the numerical values at any given point of time can be found automatically from the graph concerned. Therefore it is easy to find when the data have become nonlinear. Indeed, from some recent values of *N*(*t*), after taking the time series upside down we can try to fit a polynomial function. Currently in Italy, Spain and the United Kingdom the curve has started to *flatten*. Therefore for short term forecasting for these three countries, we need not go for either time series analysis or any other infectious disease modeling. We can easily find polynomials of a proper degree that may be used for extrapolating the values at some near future date. In the cases of these countries, a polynomial expression would be sufficient because the situation is no longer nearly exponential as the graphs concerned show.

In India, the graph shows that the spread pattern is very highly nonlinear. Indeed, it can be surmised that the pattern is nearly exponential. To observe how far this is true, we just have to see if the natural logarithm of *N*(*t*) is approximately linear. Let us suppose that

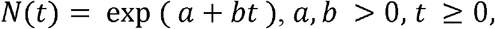

where *a* and *b* are constants. Then for

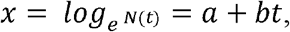

the first order differences Δ*x* of *x* would be constant for equidistant *t*. Now in reality *b* cannot be a constant for the COVID spread data because if that has to be true then the pandemic would continue spreading exponentially forever. It has been seen that in other parts of the world the pandemic has started to subside at different dates, and expectedly in India also it cannot just continue to grow exponentially, it can only be nearly exponential at present. Therefore *b* cannot be an absolute constant. It has to be time dependent.

Without going into a statistical time series analysis involving random errors, we may now use the following numerical procedure to estimate *b* looking at the time series data upside down for a recent and short period. The values of Δ*x* can be averaged for a short period, and that would give us an estimate of *b* for a short period. This was worked out in [10] for the Indian COVID19 spread data published by Worldometer.info from May 25 to May 31. In Table-1, we have shown how correct the forecasts can be if we use the numerical method explained in [10]. With May 31 as base date and with Δ*x* = 0.045, the average of Δ*x* for the period May 25 to May 31, forecasts were made for the period June 1 to June 7.

**Table-1:** Forecasts of Total Number of Cases in India from June 1 to June 7.

| Dates | Forecasts | Observed Values | Overestimation |
| --- | --- | --- | --- |
| June 1 | 199382 | 198370 | 0.51 % |
| June 2 | 208559 | 207191 | 0.66 % |
| June 3 | 218159 | 216824 | 0.61 % |
| June 4 | 228200 | 226713 | 0.65 % |
| June 5 | 238704 | 236184 | 1.06 % |
| June 6 | 249691 | 246622 | 1.24 % |
| June 7 | 261183 | 257486 | 1.43 % |

We can see that the error margins were observably low, and that the margins become larger as *t* increases. Precisely this is what we were expecting. We have expected slight overestimation which we have seen. Our standpoint is that if looking at the time series data upside down and using just a few values from the series can help us to forecast in such a good way, then we can successfully utilize our procedure for short term forecasting in this kind of a time series dealing with pandemic data. This approach would of course be of use only after the spread pattern takes an observable shape, and therefore at the initial stage this method would not lead to good results.

## RESULTS

It has been observed that the spread pattern in India is nearly exponential already. Hence for forecasting, we shall use the expression

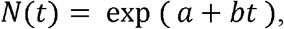

where the constant *a* would be the value of *x* on the base date June 9, and the parameter *b* would be replaced by its estimate, the average value of Δ*x* for the period from June 1 to June 9.

In Table-2, we have shown the observed values of *N*(*t*) and *x* for India from June 9 to June 1 downwards. The average value of Δ*x* for the values of *x* for these 9 days was found to be 0.04063. For the purpose of extrapolation, we shall take Δ*x* = 0.040. Therefore with June 9 as the base date, we can now extrapolate the values of the total number *N*(*t*) of cases using the formula

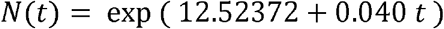

where 12.52372 is the value of *x* the base date June 9, and *t* = 1, 2, and so on for Junec10, June 11, and so on.

**Table-2:** Values of *N*(*t*) and Δ*x* for India from June 1 to June 9 Downwards.

| Dates | $N(t)$ | $X$ | $\Delta x$ |
| --- | --- | --- | --- |
| June 9 | 274780 | 12.52372 | 0.03274 |
| June 8 | 265928 | 12.49098 | 0.03226 |
| June 7 | 257486 | 12.45872 | 0.04310 |
| June 6 | 246622 | 12.41561 | 0.04324 |
| June 5 | 236184 | 12.37236 | 0.04092 |
| June 4 | 226713 | 12.33144 | 0.04459 |
| June 3 | 216824 | 12.28684 | 0.04544 |
| June 2 | 207191 | 12.24139 | 0.04350 |
| June 1 | 198370 | 12.19788 | 0.03991 |

**Table-3:** Forecasts of the Total Number of Cases in India from June 10 to June 24.

| **Dates** | **Forecasts** |
| --- | --- |
| June 10 | 285994 |
| June 11 | 297666 |
| June 12 | 309814 |
| June 13 | 322457 |
| June 14 | 335617 |
| June 15 | 349314 |
| June 16 | 363570 |
| June 17 | 378407 |
| June 18 | 393850 |
| June 19 | 409924 |
| June 20 | 426653 |
| June 21 | 444065 |
| June 22 | 462188 |
| June 23 | 481050 |
| June 24 | 500682 |

As per our forecasts, the total number of cases in India would cross 300,000 on June 12. By around June 19, it would cross 400,000, and by around June 24, it would cross 500,000. These forecasts would be slight overestimations, but in terms of percentages of the actual observations it would not be too different from the realities. Finally, if it so happens that due to natural reasons or otherwise, the situation improves as it did in Italy and Spain, the forecasts would not work because they have been made with an assumption that in India the growth would continue to be nearly exponential up to June 24.

In Table-1, we can observe that the values of Δ*x* for June 9 and June 8 were 0.03274 and 0.03226 respectively. In other words, there has been a declination of growth of the total number of cases on these two days although the average value of Δ*x* for these 9 days has been found to be 0.040. Therefore while forecasting as we have used June 9 as the base date, for the first one or two days there may actually be very slight underestimations. Thereafter the forecasts would be slight overestimations as observed in the forecasts from June 1 to June 7.

It was observed in [10] that from May 11 to May 24, the average value of Δ*x* was 0.050, and from May 25 to May 31, it was 0.045. We have observed that from June 1 to June 9, the average was 0.040. In other words, the average of Δ*x* is decreasing with time. This is the reason why we cannot go for long term forecasting, because within the period of our forecasts the average value of Δ*x* may reduce further making our forecasts overestimations thereby.

These forecasts would not be valid if the growth suddenly starts declining naturally or otherwise. If for example, lockdown is imposed in India once again, there may be declination in the total number of cases, and in that kind of a situation our forecasts might go wrong.

This method may actually lead to slightly overestimated values if we consider the entire world as a whole. When the pandemic had only just started in some countries, in certain others it was at its peak. In some countries the growth has started to retard now while in some others it is still growing at an alarming rate. For this heterogeneity, this method of forecasting of the spread taking the entire world as a whole may not give perfect forecasts, but it would certainly give some workable short term forecasts that would be slight overestimates. In any country, heterogeneity may not be too high, and hence for any specific country such a numerical approach would be useful for short term forecasts.

## CONCLUSIONS

Taking a few recent values of a time series that consists of data of a pandemic spread, we can go for short term forecasting using numerical analytical methods. Because such forecasts are not stochastic in nature, we cannot make statements in terms of confidence intervals. However, this method of forecasting does return values that can be very close to the realities. In this article, we have made forecasts of the total number of COVID-19 cases in India for a fortnight, from June 10 to June 24. The forecasts were prepared after observing that the spread pattern in India is very nearly exponential. These forecasts might go wrong if the spread retards naturally or if some measures such as re-imposition of countrywide lockdown within the period of the forecasts are taken.

## Data Availability

The data were taken from Worldometers.info .

https://www.worldometers.info/coronavirus/coronavirus-cases/

